# Socioeconomic and gender disparities in tobacco smoking among Jamaican adults from a national health survey

**DOI:** 10.1101/2021.01.04.21249206

**Authors:** Nadia R Bennett, Trevor S Ferguson, Novie O Younger-Coleman, Damian K Francis, Simon G Anderson, E. Nigel Harris, Marlene Y MacLeish, Rainford J Wilks, U.S. Caribbean Alliance for Health Disparities Research Group (USCAHDR):

## Abstract

**Objectives:** Little is known of socioeconomic and gender disparities in tobacco use in the Caribbean. We evaluated education and occupation disparities in tobacco smoking prevalence in Jamaica.

**Methods:** Data on tobacco smoking, education attainment and usual occupation in adults 25-74 years in a national survey collected between 2007 and 2008 was analyzed. Using post stratification survey weights, Poisson regression models estimated sex-specific, age-adjusted prevalence estimates, prevalence differences and prevalence ratios.

**Results:** Analyses included 2299 participants (696 men, 1603 women), mean age 43 years. Current smoking prevalence was 26% in men and 8% in women (p<0.001). Among men, age adjusted prevalence of current smoking was highest in primary education (36.5%) and lowest in the post-secondary education groups (10.2%), (p= 0.003). Among women, prevalence was highest in junior secondary education (10.2%) and lowest in primary education groups (4.7%), (p = 0.014). Among men, for education, age-adjusted prevalence ratios for current smoking ranged from 2.6 to 3.6 using post-secondary education as the reference category (p<0.05). For occupation, age-adjusted prevalence ratios ranged from 1.7 to 4.1 using professionals and managers as the reference category. Among women, using the same reference categories age-adjusted prevalence ratios for education ranged from 1.4 to 2.2 and for occupation 0.6 to 2.2, neither were statistically significant.

**Conclusion:** In Jamaica, there are socioeconomic disparities in current tobacco smoking among men, where it is inversely associated with education attainment and occupation but in women is less clear. These findings suggest interventions to reduce smoking should consider these disparities.

---

Tobacco use is the second leading contributor to the global burden of disease and the leading cause of preventable death worldwide (1, 2). Approximately 80% of the 1.4 billion smokers live in lower and middle income countries (LMIC), where the burden of tobacco related death is the greatest (3). Half of current tobacco users will die from a tobacco related disease (4). The global prevalence of current tobacco use in adults was approximately 22% in 2012 and had wide regional variation(5). In Jamaica, the overall estimated prevalence of current tobacco use was 17.7% in 2000-2001 and 14.5% in 2007-2008 (6, 7).

Socioeconomic factors such as relative income and education are important contributors to the aetiology of a wide range of health outcomes. There is consistent evidence for a strong association between the prevalence of non-communicable diseases (NCD) and social factors such as education level, occupation and gender (8, 9). In high income countries, the risk of having a NCD is greater in those of lower socioeconomic status (SES), however, in low- and middle-income countries (LMICs), this inverse relationship between SES and the presence of NCDs is inconsistently observed (8, 10). There are several individual and societal factors such as level of education, type of occupation, public health policy, social and cultural norms which variably contribute to the disparities and may change the relationships observed (8, 11, 12).

Socioeconomic disparities in tobacco smoking have been previously reported (13, 14). Data from the world health surveys shows that among men, the prevalence of tobacco smoking was twice as high among those with no formal schooling when compared to those who had completed college education or above (13). There are limited data on disparities in tobacco smoking in Caribbean origin populations. Figueroa and colleagues reported that lower educational status was significantly associated with smoking cigarettes and marijuana among men in Jamaica (15), while Stringhini and colleagues also found that smoking prevalence was higher among persons with lower education in urban Jamaica (14).

This study aimed to evaluate the association between socioeconomic status (SES), using education and occupation as indicators of status, and prevalence of current tobacco smoking using data from the Jamaica Health and Lifestyle Survey 2007-2008 (JHLS-II). We hypothesized that lower educational attainment and occupation category were associated with a higher prevalence of current tobacco smoking. We also aimed to evaluate whether gender acts as a risk modifier in the association between SES and smoking.

## Methods

We analyzed data from a national health examination survey (The Jamaica Health and Lifestyle Survey) conducted in Jamaica between November 2007 and February 2008. This survey sought to determine the health status, nutritional habits, lifestyle and behaviour in a nationally representative sample of Jamaican adults(7).

Using a multistage sampling design, 2848 Jamaicans between 15-74 years were selected for inclusion in the survey. Enumeration district (ED) served as the primary sampling units (PSU) and were randomly selected, using a probability propionate to size sampling strategy. Households within each PSU were systematically selected, using a random starting point. The sampling interval varied between EDs and was determined by the number of households in the ED, in order to recruit 20 participants within each ED. One participant from each household was selected using the Kish selection method (16). Additional details of the study design were described and published in a full report (7).

For this study, participants eligible for inclusion were restricted to individuals 25-74 years old to include those with completed post-secondary education. Ethical approval was obtained from the University of the West Indies, Faculty of Medical Sciences Ethics Committee, and the Ministry of Health Ethics Committee. Written, informed consent was obtained from each participant prior to data collection.

### Variables: Measurement and Definitions

An interviewer-administered questionnaire was used to collect socio-demographic data, information on tobacco use, other health behaviours, health status and health care practices. Study participants reported smoking history and current smoking by answering positively to the following questions: (1) *“Do you currently smoke any form of tobacco (cigarettes, beady etc*.*)?” and (2) “Did you ever smoke any form of tobacco (cigarettes, beady, etc*.*)?”* Study participants who responded negatively to both questions were classified as never smokers, those who answered yes to question 1 as current smokers and those who answered yes as former smokers. For these analyses we used a dichotomous variable based on those who answered yes to question 1.

Health disparity defined by educational attainment and occupation category were the exposures of interest. The level of educational attainment was divided into 4 categories; 1) primary or lower (up to grade 6); 2) junior secondary (up to grade 9); 3) full secondary (at least grade 11); and 4) post-secondary; (vocational training, college or university). Occupation category was defined by the Jamaica Standard Occupational Classification 1991 (JSOC) which divided occupation into 10 groups which were subsequently collapsed in four categories for analysis: 1) Professionals & Managers (JSOC 1-3), 2) Office & Service Workers (JSOC 4-5, 10), 3) Trade Workers & Farmers (JSOC 6-8), and 4) Elementary Occupations (JSOC 9)(17). We added one other category for those who were, students, retired housewives or currently unemployed. Individuals whose occupation did not fit into any pre-defined category and non-responders to the question on occupation were excluded from the analyses.

### Statistical analyses

Data were analysed using Stata 12.1 statistical software (Stata Corp., College Station Texas). To account for multi-stage survey design, post-stratification survey weights were applied to the data using the survey commands in Stata.

We performed descriptive analyses yielding means or proportions for demographic variables, tobacco use, education level and occupation categories as appropriate. We calculated crude and sex-specific estimates of the prevalence of current tobacco smoking within and across education, occupation, and age categories. Poisson regression models were used to calculate age-adjusted prevalence, prevalence difference and prevalence ratios. Prevalence ratios were used for these analyses given that it has been shown to be a good measure of effect where the prevalence of the outcome is high (18, 19). Individual models were created for each of education and occupation and post-estimation commands used to derive adjusted estimates. Interactions between sex and age-group were tested in the regression models. Any statistically significant associations were included in the final models. There was evidence for sex interaction in the association between SES (education and occupation) and smoking, therefore we present sex-specific estimates for prevalence and multivariable models. Age-adjusted estimates were obtained from models which included the interaction terms. Analyses were limited to participants 25-74 years with complete data on education and smoking status.

## Results

Analysis included 2299 persons (696 males, 1603 females) with a mean age of 42.9 rears (range 24-74) years. Descriptive analyses are shown in Table 1. Overall, the prevalence of current smoking was 16.4% and was higher among men (25.8% vs. 7.8%, p <0.001) compared to women. There was evidence of significant sex differences in education attainment (p<0.05) and occupation category (p<0.001). A higher proportion of females (63.3%) compared to males (57.5%) had completed full secondary education or post-secondary education; however, a higher proportion of men were in the professionals and managers category (18.5 vs. 9.8%).

**Table 1:** Summary statistics^1^ for characteristics of 25-74 year old participants from the Jamaica Health and lifestyle survey 2007-2008 (JHLS-II)

|  | Male<br>(n=696) | Female<br>(n=1603) | Total<br>(n=2299) |
| --- | --- | --- | --- |
| **Age years** mean (SE) | 42.9 (0.11) | 42.8 (0.09) | 42.9 (0.07) |
| **Age groups, % (n)** |  |  |  |
| 25-39 years | 45.6 (244) | 46.0 (620) | 45.8 (864) |
| 40-59 years | 41.2 (293) | 40.7 (698) | 40.9 (991) |
| >60 years | 13.2 (159) | 13.3 (285) | 13.2 (444) |
| **Education levels\*, % (n)** |  |  |  |
| Post-secondary | 11.7 (64) | 9.6 (142) | 10.6 (206) |
| Full Secondary | 45.7 (276) | 53.6 (761) | 49.8 (1037) |
| Junior Secondary | 27.3 (206) | 23.2 (419) | 25.2 (625) |
| Primary or lower | 15.3 (150) | 13.6 (281) | 14.4 (431) |
| **Occupation<sup>2</sup> category \*\*\*, % (n)** |  |  |  |
| Professionals & Managers | 19.7 (112) | 11.0 (141) | 15.2 (253) |
| Office & service workers | 22.8 (131) | 47.0 (626) | 35.3 (757) |
| Trade workers & Farmers | 47.9 (333) | 10.3 (171) | 28.5 (504) |
| Elementary occupations | 7.9 (47) | 17.3 (231) | 12.8 (278) |
| Other | 1.6 (17) | 14.4 (242) | 8.2 (259) |
| **Smoking status\*\*\*, % (n)** |  |  |  |
| Never | 49.0 (319) | 81.1 (1304) | 65.7 |
| Past | 25.2 (188) | 11.1 (181) | 17.9 |
| Current | 25.8 (189) | 7.8 (118) | 16.4 |

<sup>1</sup>Proportions (%) and means were weighted for survey design to provide population based estimates.
<sup>2</sup>Estimates for occupation included 2051 participants (1411 females and 640 males)
\*p <0.05, \*\*p <0.01, \*\*\*p <0.001 for male-female differences

The distribution of education attainment and occupation category by age group and sex are shown in Table 2. Age group was strongly associated with occupation and education in both males and females (p<0.001 for all comparisons). In both males and females those >60 years had the lowest level of education. The highest proportion of men in all age groups were in the trade workers and farmers occupation category, whereas the highest proportion of women were in the office and service workers occupational group.

**Table 2:** Sex-specific distribution participants by age and socioeconomic status in JHLS-II

|  | Men |  |  | Women |  |  |
| --- | --- | --- | --- | --- | --- | --- |
|  | 25-39 years | 40-59 years | >60 years | 25-39 years | 40-59 years | >60 years |
| **Education** |  |  |  |  |  |  |
| Post-Secondary | 16.7 (34) | 8.8 (25) | 3.6 (5) | 13.7 (81) | 5.9 (48) | 6.4 (13) |
| Full Secondary | 59.1 (146) | 42.4 (115) | 9.4 (15) | 68.6 (413) | 51.5 (327) | 8.2 (21) |
| Junior Secondary | 22.3 (59) | 32.3 (99) | 28.7 (48) | 16.0 (115) | 22.9 (220) | 27.7 (84) |
| Primary or lower | 1.9 (5) | 16.5 (54) | 58.3 (91) | 1.7 (11) | 12.7 (103) | 57.7 (167) |
| **Occupation<sup>1</sup>** |  |  |  |  |  |  |
| Professional | 22.6 (49) | 19.3 (51) | 9.4 (12) | 13.3 (71) | 9.6 (56) | 7.6 (141) |
| Office | 30.4 (71) | 15.9 (38) | 17.7 (22) | 48.8 (271) | 48.7 (287) | 47.0 (626) |
| Trade | 40.0 (98) | 56.3 (169) | 49.4 (66) | 7.9 (49) | 12.9 (94) | 10.3 (171) |
| Elementary | 7.0 (16) | 8.5 (23) | 9.5 (8) | 19.4 (102) | 16.7 (102) | 17.3 (231) |
| Other | 0 (0) | 0 (0) | 13.9 (17) | 10.6 (63) | 12.1 (88) | 14.4 (242) |

Estimates for occupation included 2051 participants (1411 females and 640 males)
P <0.001 for difference in distribution of education and occupation categories by age for both men and women.

Prevalence of current smoking within education, occupation and age categories is shown in Table 3. Women smoked significantly less than men and the heterogeneity of smoking prevalence across educational and occupational categories among women was much smaller than among men (education: M 9-7-38.6; F 5.6-9.7; occupation: M 12.1-52.4; F 2.9-11.8). There were significant differences in the prevalence of smoking across education categories in men but not women. Among men prevalence of smoking was higher in those with lower education ranging from 38.6% in those with primary or lower education to 9.7% among those with post-secondary education (p=0.002). There were no statistically significant differences in the prevalence of current smoking by education among women which ranged between 5.6% for post-secondary education and 9.7% in junior secondary. Occupation category was significantly associated with current tobacco use in both men and women. Among men the prevalence ranged between 12.1% in professionals and managers and 52.4% in those with elementary occupations. In women the lowest prevalence was among trade workers and farmers (2.9%) and the highest in elementary occupations (11.8%). Age group was significantly associated with current tobacco use in men (p=0.026) but among women, failed to achieve conventional levels of statistical significance (p=0.059). In men the highest prevalence was in the >60 age group and in women it was in the 25-39 age group.

**Table 3:** Prevalence^1^ of current smoking by education, occupation, and age categories among Jamaicans aged 25-74 years old from JHLS-II

|  | Male | Female | Total |
| --- | --- | --- | --- |
| **Education** | **n = 696** | **n=1603** | **N = 2299** |
| Post-secondary | 9.7 (8) | 5.6 (7) | 7.7 (15) |
| Full Secondary | 25.1 (73) | 7.6 (59) | 15.3 (132) |
| Junior Secondary | 26.7 (56) | 9.7 (34) | 18.5 (90) |
| Primary or lower | 38.6 (52) | 6.8 (18) | 22.9 (70) |
| p-value | 0.002 | 0.431 | 0.002 |
| **Occupation** | **n = 640** | **N = 1411** | **N = 2051** |
| Professionals & Managers | 12.1 (14) | 6.8 (8) | 10.1 (22) |
| Office & service workers | 25.6 (36) | 7.3 (51) | 13.0 (87) |
| Trade workers & Farmers | 27.2 (96) | 2.9 (6) | 22.7 (102) |
| Elementary occupations | 52.4 (23) | 11.8 (21) | 24.0 (44) |
| Other | 30.1 (5) | 5.8 (13) | 8.2 (18) |
| p-value | <0.001 | 0.03 | <0.001 |
| **Age years** | **n = 696** | **n = 1603** | **N = 2299** |
| 25-39 | 20.9 (55) | 10.0 (60) | 15.2 (115) |
| 40-59 | 28.9 (83) | 6.2 (45) | 17.0 (128) |
| >60 | 33.9 (51) | 5.2 (13) | 18.8 (64) |
| p-value | 0.026 | 0.059 | 0.431 |

<sup>1</sup>Proportions (%) and means were weighted for survey design to provide population based estimates

Multivariable analyses conducted using Poisson regression models yielding age adjusted prevalence estimates, prevalence differences and prevalence ratios are shown in Table 4. As previously indicated, we present sex-specific models in light of sex interaction in the association between smoking and both education and occupation. Among men, age adjusted prevalence of smoking was highest among those with primary or lower education and lowest in those with post-secondary education. In women age adjusted prevalence was highest among those who had junior secondary education and lowest among those with post-secondary education. Using post-secondary education as the reference category there was a statistically significantly higher prevalence for current tobacco use among all other education categories for men with prevalence ratios ranging between 2.6 for full secondary to 3.6 for the primary education category. Similarly, the prevalence differences ranged between 16.1% and 26.3%. Among women the only significant difference was 5.4% for those who completed full secondary education. The findings were similar when occupation was used as a measure of SES with significant differences for men but not for women. Among men, prevalence ratios ranged between 2.2 for office and service workers and 4.1 for elementary occupations. The prevalence differences ranged between 15.0% - 40.9% for office and service workers and elementary occupations.

**Table 4:** Age-adjusted^***1***^ prevalence estimates, prevalence difference and prevalence ratiosfor education and occupation categories for males and females in the JHLS-II

| Characteristic | Male |  | Female |  |
| --- | --- | --- | --- | --- |
| Education |  |  |  |  |
| Prevalence | % (95% CI) |  | % (95% CI) |  |
| Post-Secondary | 10.2 (2.4 - 18.2) | - | 4.7 (0.8 -8.5) | - |
| Full Secondary | 26.4 (21.4 - 31.3) | - | 6.5 (4.7 -8.4) | - |
| Junior Secondary | 26.8 (19.1 - 34.5) | - | 10.2 (6.7 -13.6) | - |
| Primary or lower | 36.5 (25.7 - 47.3) | - | 9.8 (3.9 – 15) | - |
| Prevalence Ratio (PR) | PR (95% CI) | P-Value | PR (95% CI) | P-Value |
| Post-Secondary | 1 |  | 1 |  |
| Full Secondary | 2.6 (1.2 -5.6 ) | 0.018 | 1.4 (0.6 -3.2) | 0.439 |
| Junior Secondary | 2.6 (1.1 -6.1) | 0.026 | 2.2 (1.0 – 4.8) | 0.057 |
| Primary or lower | 3.6 (1.5 -8.3) | 0.003 | 2.1 (0.9 -4.9) | 0.1 |
| Prevalence difference (PD) | PD (95% CI) | P-Value | PD (95% CI) | P-Value |
| Post-Secondary | 0 |  | 0 |  |
| Full Secondary | 16.1 (7.2 - 2.0) | <0.001 | 1.8 (-2.2 - 5.9) | 0.375 |
| Junior Secondary | 16.6 (5.2 - 27.9) | 0.004 | 5.4 (1.1 - 9.8) | 0.014 |
| Primary or lower | 26.3 (12.6 – 39.9) | <0.001 | 5.1 (-1.0 - 9.8) | 0.1 |
| Occupation<sup>2</sup> |  |  |  |  |
| Prevalence | % (95% CI) |  | % (95% CI) | - |
| Professionals and Managers | 13.0 (6.3 -19.7) | - | 5.6 (0.9- 10.2) | - |
| Office and Service Workers | 28.1 (19.1 -37.0) | - | 7.5 (5.0 -10.0) | - |
| Trade Workers and Farmers | 27.7 (21.9 -33.4) | - | 3.4 (0.4-6.4) | - |
| Elementary Occupations | 54.0 (36.2 -71.7) | - | 12.4 (7.0 -17.7) | - |
| Other/Unemployed | 22.2 (3.0 - 41.4) | - | 5.2(0.4 -10.0) | - |
| Prevalence Ratio (PR) | PR (95% CI) | P-Value | PR (95% CI) | P-Value |
| Professionals and Managers | 1 |  | 1 |  |
| Office and Service Workers | 2.2 (1.2-3.8) | 0.007 | 1.3 (0.6 -3.2) | 0.503 |
| Trade Workers and Farmers | 2.1 (1.2 -3.8) | 0.010 | 0.6 (0.2 -2.1) | 0.439 |
| Elementary Occupations | 4.1 (2.2 -7.8) | <0.001 | 2.2 (0.83 -5.9) | 0.112 |
| Other/Unemployed | 1.7 (0.6 -4.7) | 0.301 | 0.9 (0.3 -2.9) | 0.907 |
| Prevalence Difference (PD) | PD (95% CI) | P-Value | PD (95% CI) | P-Value |
| Professionals and Managers | 0 |  | 0 |  |
| Office and Service Workers | 15.0 (4.9 -25.1) | 0.004 | 1.9 (- 3.1 -7.0) | 0.456 |
| Trade Workers and Farmers | 14.6 (5.5 -23.8) | 0.002 | -2.2 (-7.8 -3.4) | 0.453 |
| Elementary Occupations | 40.9 (21.5 -60.4) | <0.001 | 6.8 (-0.6 -14.2) | 0.073 |
| Other/Unemployed | 9.1 (-11.2 – 29.3) | 0.377 | -0.3 (-0.6 – 5.7) | 0.904 |

<sup>1</sup>Estimates were derived from Poisson regression models adjusted for age as categorical variable. Separate models were created for occupation and education. Age-adjusted prevalence estimates and prevalence difference were derived from the models using post estimation commands.
<sup>2</sup>Estimates for occupation included 640 men and 1183 women. Estimates for women excluded women in the 60 -74 years old age group and some occupation categories had no current smokers.

## Discussion

In this study, one in six adults were current tobacco users, with a three-fold higher prevalence among men compared to women. There were significant disparities in the prevalence of current tobacco use by education and occupation categories among adult men in Jamaica. Among men, the prevalence of current smoking was four-fold higher among those in the lowest education or occupation categories compared to those in the highest categories. In women, the prevalence of current tobacco use was higher among those in lower education or occupation categories, but the observed differences did not achieve statistical significance.

The findings in this study were generally consistent with the published literature with a clear negative gradient for men in most countries, but a more varied pattern of negative and positive gradients among women (13, 20, 21). In most high income countries and some lower and middle income countries (LMICS), the prevalence of tobacco use overall is higher among persons with less education and lower socioeconomic status (22-24). However, in some middle income countries it has been observed that the prevalence of tobacco smoking increases as educational attainment increases particularly among older women (20). In a study from Malawi and Zambia, the author reported similar findings to ours, where there was a strong and statistically significant negative gradient among men for both education and occupation, weaker and mostly non-significant associations for occupation among women (21). Taken together these studies suggest that socioeconomic health disparities in tobacco smoking is more clearly defined among men in developing countries, where smoking is significantly more prevalent in men than in women and the tendency for higher prevalence among lower SES groups is more marked and consistently statistically significant.

Based on these findings it is plausible to expect that the negative impact of cigarette smoking in developing countries may be greatest among men with lower SES. These persons may also be the least likely use available health services and therefore the risk of adverse outcomes will be quite high. Additionally, men in lower SES categories often have large numbers of dependents, so the consequences of adverse outcomes have much greater impact on their families and communities. Interventions designed to reduce smoking in the Jamaica and similar developing countries will therefore need to take into account these sex and SES differences, with special programmes targeting highest risk groups, particularly lower SES men. Further studies will need to explore the sociocultural factors that result in high prevalence smoking in these groups in order to design culturally appropriate interventions.

A major strength of this study is the use of a relatively large nationally representative sample with a 98% response rate. Additionally, we used survey weights to adjust for deviations of the sample from population characteristics. We are therefore confident that the findings can be generalized to the general Jamaican population. Given that social and cultural characteristics in Jamaica are generally similar to that seen in other English-speaking Caribbean countries, the findings will therefore be relevant to these counties and others with similar sociocultural characteristics. The findings should provide the basis for targeted intervention strategies in countries of similar stage of social and economic development.

The study was limited by its restriction to current tobacco use and the absence of quantitative estimates of cigarettes smoking as well as duration of tobacco use. We were therefore unable to evaluate whether there was a dose response relationship in the association between SES and tobacco smoking. Additionally, we found that there were small numbers in some sub-groups particularly among women, which may have resulted in unstable estimates. We also found that the occupations reported by some of the participants did not fit into the classification scheme used and as such had to be excluded from the analyses for occupation. The occupational analyses was limited by the loss of 11% of participants who did not fit into any of the categories. This as an a priori decision and its impact is difficult to estimate.

## Conclusion

The heterogeneity of smoking prevalence among women by educational and occupational categories was remarkably smaller than among men and there was significant interactions between sex and education/occupation mandated sex-specific modeling. We have found significant socioeconomic disparities in tobacco smoking among men in Jamaica, but weaker and less clear associations among women. The high smoking rates among men in the lower SES categories is of major public health concern and is likely to present a major burden on the nation’s health resources. Further research should seek to identify the social and cultural drivers of these disparities and design interventions to reduce tobacco use, with special emphasis on lower SES men.

## Data Availability

The datasets analyzed during the current study available from the corresponding author on reasonable request.

## Acknowledgments

The authors wish to thank the respondents who participated in the survey and the training and field staff, who worked to ensure the timely completion of the survey. Thanks also to other investigators involved in the Jamaica Health and Lifestyle Survey 2007-2008 (JHLS-II) and investigators from the United States Caribbean Alliance for Health Disparities Research (USCADDR) for their contribution to this project.

